# Linear epitope landscape of SARS-CoV-2 Spike protein constructed from 1,051 COVID-19 patients

**DOI:** 10.1101/2020.07.13.20152587

**Authors:** Yang Li, Ming-liang Ma, Qing Lei, Feng Wang, Dan-yun Lai, Hongyan Hou, Zhao-wei Xu, Bo Zhang, Hong Chen, Caizheng Yu, Jun-biao Xue, Yun-xiao Zheng, Xue-ning Wang, He-wei Jiang, Hai-nan Zhang, Huan Qi, Shu-juan Guo, Yandi Zhang, Xiaosong Lin, Zongjie Yao, Jiaoxiang Wu, Huiming Sheng, Ziyong Sun, Xionglin Fan, Sheng-ce Tao

**Affiliations:** Shanghai Center for Systems Biomedicine, Key Laboratory of Systems Biomedicine (Ministry of Education), Shanghai Jiao Tong University, Shanghai, China; Department of Pathogen Biology, School of Basic Medicine, Tongji Medical College, Huazhong University of Science and Technology, Wuhan, China; Department of Clinical Laboratory, Tongji Hospital, Tongji Medical College, Huazhong University of Science and Technology, Wuhan, China; Department of Public Health, Tongji Hospital, Tongji Medical College, Huazhong University of Science and Technology, Wuhan, China; Tongren Hospital, Shanghai Jiao Tong University School of Medicine, Shanghai, China

## Abstract

Neutralization antibodies and vaccines for treating COVID-19 are desperately needed. For precise development of antibodies and vaccines, the key is to understand which part of SARS-CoV-2 Spike protein is highly immunogenic on a systematic way. We generate a linear epitope landscape of Spike protein by analyzing serum IgG response of 1,051 COVID-19 patients with a peptide microarray. We reveal two regions that rich of linear epitopes, *i.e*., CTD and a region close to the S2’ cleavage site and fusion peptide. Unexpectedly, we find RBD is lack of linear epitope. Besides 3 moderate immunogenic peptides from RBD, 16 highly immunogenic peptides from other regions of Spike protein are determined. These peptides could serve as the base for precise development of antibodies and vaccines for COVID-19 on a systematic level.

**One sentence summary:** A linear epitope landscape of SARS-CoV-2 Spike protein is generated by analyzing serum IgG response of 1,051 COVID-19 patients.

---

COVID-19 is caused by SARS-CoV-2 (*1, 2*) and is a global pandemic. By July 12, 2020, 12,728,966 cases were diagnosed and 565,351 lives were claimed (https://coronavirus.jhu.edu/map.html) (*3*). Highly effective neutralization (prophylactic or therapeutic) antibodies and vaccines are pressingly needed. The genome of SARS-CoV-2 encodes 27 proteins, among them, Spike protein plays a central role for the binding and entry of the virus to the host cell. Spike protein is cleaved into S1 and S2 at furin and S2’ sites by specific proteases (*4*). It is highly glycosylated with 21 N-glycosylation sites (*5*). Spike protein and more specific, the RBD domain is currently the most-focused target for the development of COVID-19 neutralization antibodies and vaccines (*6*–*11*). However, except RBD, other parts on Spike protein may also elicit neutralization antibodies (*12*–*16*). In addition, antibody dependent enhancement (ADE) is an unneglectable concern for the development of neutralization antibody and vaccine for SARS-CoV-2 (*17, 18*). To facilitate the precise development of neutralization antibodies and vaccines, it is necessary to understand which part of Spike protein is highly immunogenic on a systematic way, by analyzing samples collected from a large cohort of COVID-19 patients.

## Results

### The IgG linear epitope landscape of SARS-CoV-2 Spike protein

To reveal the immunogenic linear epitopes of Spike protein, a peptide microarray with full coverage of Spike protein was updated from an original version (*14*) (**Fig. S1**). Sera were collected from two groups, *i.e*., 1,051 COVID-19 patients and 528 controls (**Table S1**), and analyzed individually on the peptide microarray (**Table S2)**. By plotting the signal intensities of all the samples against each peptide, a linear epitope landscape was constructed, for better overview, the landscape is aligned to the sequence of Spike protein (**Figure 1**). To assure the specificity, all the control samples (**Table S1**) were also analyzed on the peptide microarray. Almost all the peptides were negative for all the control samples, while significant bindings were observed for many of the peptides when probed with COVID-19 sera. This indicate that the positive bindings are SARS-CoV-2 specific. It was speculated that the N-glycosylation may interfere with antibody responses (*19*). However, our results clearly showed that the distribution of the linear epitopes is not related to N-glycosylation, if any, subtle.

**Figure 1.**
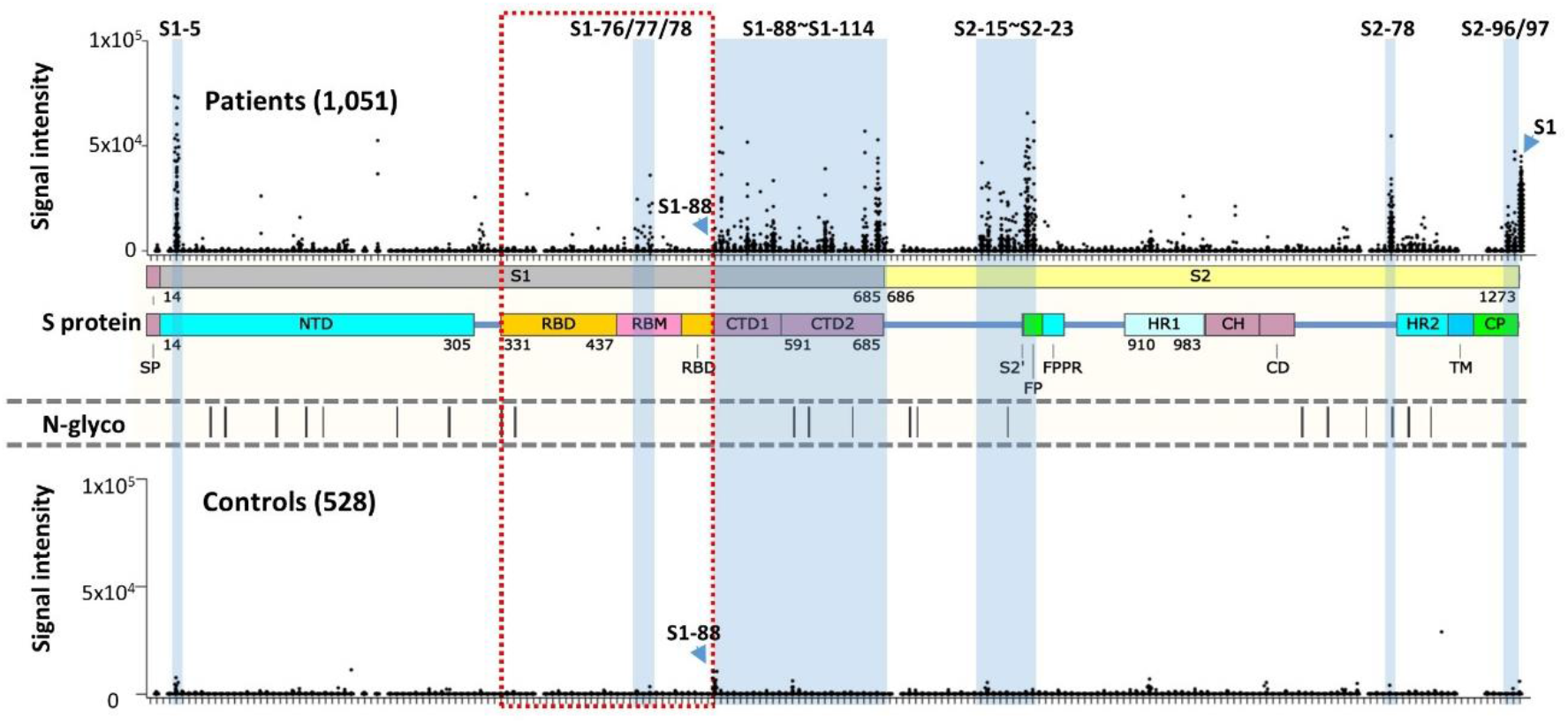
The IgG linear epitope landscape of SARS-CoV-2 Spike protein. The signal intensities of 1,051 COVID-19 sera against 197 peptides were obtained by using the peptide microarray. The peptides are listed in X-axis and aligned to the corresponding locations on Spike protein. As a control, the signal intensities of S1 protein were also presented. The missing spots are peptides either could not be synthesized or failed for BSA conjugation (see **Table S2** for details). A cohort of 528 controls sera were also analyzed on the microarray. In addition, the known N-glycosylation sites (N-glyco) were aligned with Spike protein. The peptides or regions with significant binding were marked blue. Peptide S1-88 was specifically labeled because significant bindings were also observed for the controls. SP: signaling peptide; NTD: N-terminal domain; RBD: receptor biding domain; RBM: receptor binding motif; CTD1: C-terminal domain 1; CTD2: C-terminal domain 2; S2’: protease cleavage site; FP: fusion peptide; FPPR: fusion peptide proximal region; HR1: heptad repeat 1; HR2: heptad repeat 2; CH: center helix; CD: connector domain; TM: trans-membrane; CP: cytoplasmic.

To call the highly immunogenic peptides, the criteria were set as the average_signal intensity is larger than 3*Cutoff2, and the response frequency is larger than 10% (see methods for the definitions). A total of 16 peptides were obtained, surprisingly, all of them are outside of RBD. Because the significance of RBD, we lowered the criteria, *i. e*., the response frequency is larger than 1%, while keep the average_signal intensity larger than 3*Cutoff2, 3 consecutive peptides of moderate immunogenicity, *i. e*, S1-76, S1-77 and S1-78 were selected (**Table S3)**. Interestingly, all these peptides locate within RBM (receptor binding motif), the binding interface of Spike protein and ACE2.

While a few of these immunogenic peptides are dispersed on Spike protein, there are two linear epitope “hot” regions could be immediately recognized, aa525-685 and aa770-829, one of which is CTD (C-terminal domain) and another covers the S2’ cleavage site and the fusion peptide (FP). For the immunogenic peptides (**Table S3)**, the solubility (*20*) and pI range from −1.97 to 1.06, and 3.01 to 11.16, respectively. This indicates the overall immunogenicity of Spike protein is not correlated to solubility and pI. There are several SARS-CoV-2 epitope related studies involving small sample sets (*21, 22*). Our immunogenic epitopes are partially consistent with these studies (**Table S3**). A relative high consistency was observed between our data and ReScan, a phage display based strategy (*23, 24*).

It is known that the immune response may be related to some key clinical parameters, such as gender, disease severity, age and the final outcome (*25, 26*). To test whether the linear epitope landscape correlates with these parameters, we divided the linear landscape (**Fig. 1A**) according to these parameters, *i.e*., male vs. female for gender (**Fig. S2**.), mild vs. severe for severity (**Fig. S3**), >60 vs. <60 for age (**Fig. S4**) and recovered vs. death for final outcome (**Fig. S5**). Unexpectedly, overall, no obvious difference was observed. These suggest that the linear epitope landscape is highly robust and consistent among COVID-19 patients.

To further illustrate the location and distribution of the 19 immunogenic peptides (**Table S3**), we mapped them to the 3D structure of Spike protein (*27*) (**Fig. S6A** and **S6B**). It is clear that most of these peptides locate on the surface of Spike protein, which is consistent to the common notion (*28*). However, S1-111, S2-15 and S2-16 are not on the surface of the trimeric Spike protein, but on the surface of the monomer. A plausible explanation is that the monomer Spike protein could be exposed to the immune system at a certain yet undiscovered stage.

### RBD is lack of linear epitope while highly immunogenic

It is well known that RBD is highly immunogenic (*26, 29*). However, according to our selection criteria, no highly immunogenic peptide was obtained from RBD. Only when we lowered the selection criteria, 3 peptides were selected, *i.e*., S1-76, S1-77 and S1-78 (**Fig. 2A**). When RBD is compared to other regions of Spike protein, it is obvious that RBD is very poor of linear epitopes. This seems contradictory to the knowledge that RBD is highly immunogenic. It is possible that most of the epitopes of RBD region are conformational. To test this possibility, we collected a set of 9 high affinity monoclonal antibodies of RBD or Spike protein (see methods). These antibodies were obtained through memory B cell isolation from COVID-19 recovered patients (*30*). We analyzed these antibodies individually on the Spike protein peptide microarray (*14*) (**Fig. 2B**). Among these antibodies, 414-1 has the highest affinity (2.96 nM) to RBD, as expected, strong bindings were observed for both S1 protein and RBD, however, negative signals were obtained for all the peptides, including the RBD peptides from aa331-524. Also, no peptide bindings were observed for the rest of the RBD specific antibodies (data not shown). For antibody 414-4, strong binding was obtained for S1 protein but not RBD. Interestingly, 414-4 binds S1-97 with high affinity, indicating the epitope that 414-4 recognized is around aa577-588. Actually, the 3 immunogenic peptides, *i. e*., S1-76, S1-77 and S1-78 were also identified in other related studies (*22, 31*). These three peptides are consecutive and locate in RBM region, which are at least partially overlapped with or close to the binding epitopes of a variety of SARS-CoV-2 neutralization antibodies, *e.g*., B38 (*6*), CB6 (*32*) and P2B-2F6 (*33*). To further illustrate the locations of these peptides, we mapped them to the 3D structure of Spike protein of both the open state and the closed state (**Fig. 2C**). In closed state, aa455-465 of S1-76/77/78 locate at the contact area among the three monomers and probably is difficult to be accessed, aa452-454 and 473-474 form as β-strand, and are covered but could be accessed from both sides, while only aa466-472 are exposed and present as flexible sequence (**Fig. 2C** and **Fig. S7**). In the open state of Spike protein, all residues of S1-76/77/78 are exposed and highly accessible (**Fig. S7**). To further analyze the immunogenicity of S1-76/77/78, neutralization antibody-RBD complexes are used (**Fig. S8**), the antibodies are CB6 (*32*), P2B-2F6 (*33*), BD23 (*7*), CR3022 (*8*) and S309 (*34*). Among these structures, CB6 and P2B-2F6 interact directly with residues within S1-76/77/78. For the CB6-RBD complex (**Fig. S8D**), there are several residues within S1-76/77/78, which have direct interactions with the antibody, *i. e*., Y453, L455, F456, R457, K458, S459, N460, Y473 and Q474. While for the P2B-2F6-RBD complex (**Fig. 8E**), the only residue which has direct interaction with the antibody is L452.

**Figure 2.**
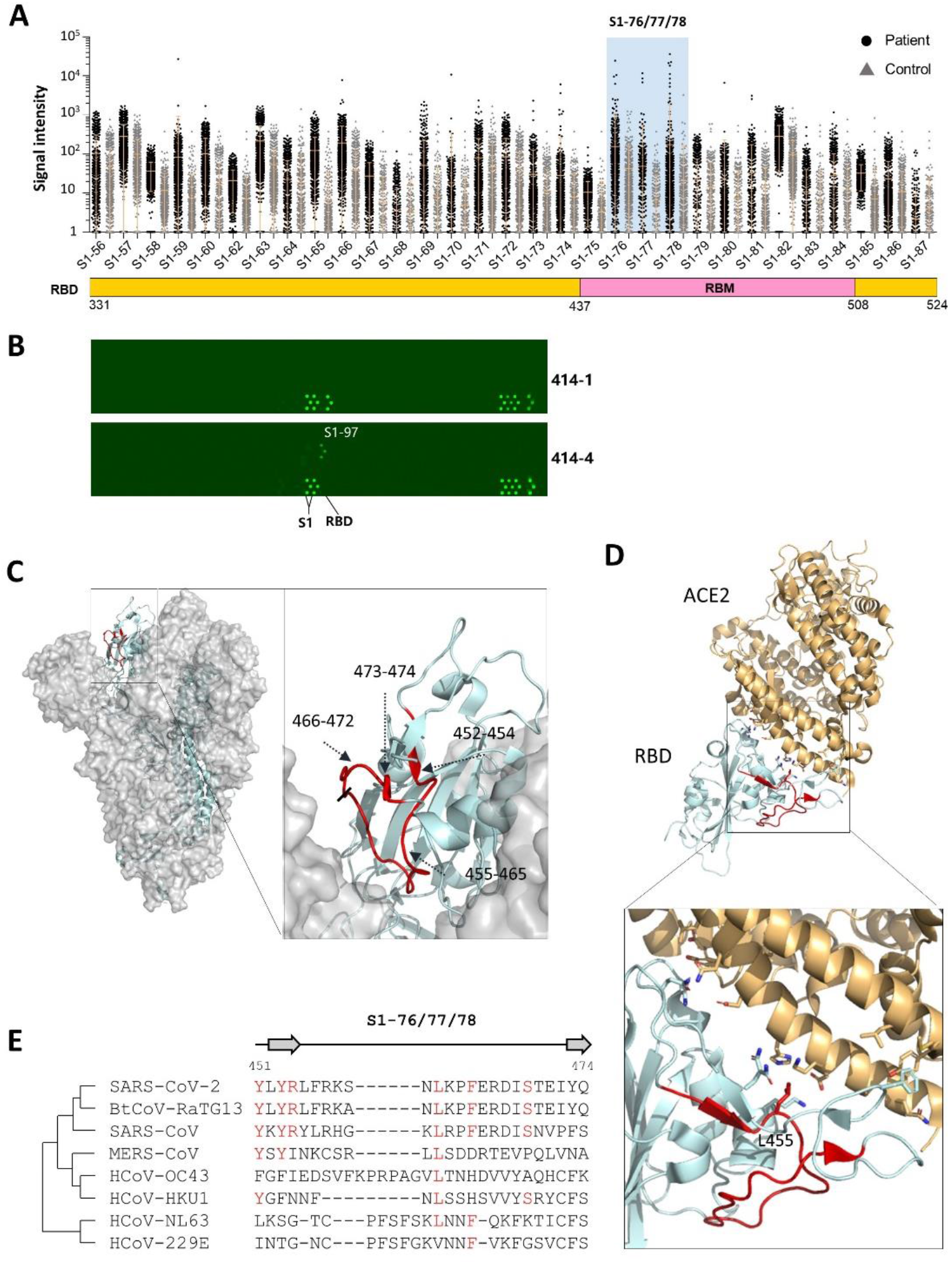
RBD is lack of highly immunogenic linear epitope. **A**. RBD region of the linear epitope landscape; **B**. The peptide microarray results of two Spike protein specific monoclonal human antibodies (from Active motif Co. Ltd.), one (414-1) is specific for RBD and the other (414-4) is not. **C**. The detailed structures of the significant peptides (S1-76/77/78, aa451-474, red) on RBD of the closed state Spike protein trimer, the side view (PDB ID: 6×6P); **D**. The locations of the significant peptides (S1-76/77/78, aa451-474, red) on the co-crystal structure of RBD and ACE2 (PDB ID: 6M0J). **E**. The homology analysis of the significant peptides among the 7 known human coronaviruses and the Bat coronavirus BtCoV-RaTG13, which is highly homologous to SARS-CoV-2, the amino acids with consistencies >=50% among the 8 coronaviruses are marked red, the loop and β-strand region are shown as line and arrow above the sequences, respectively.

Broad neutralization antibody and vaccine effective for SARS-CoV-2 and other human coronaviruses are of high interest (*35*). We performed the homology analysis for S1-76/77/78 among SARS-CoV-2, the other 6 human coronaviruses, and bat coronavirus BtCoV-RaTG13 (*1*). High homologies were observed for all these 3 peptides among SARS-Cov-2, SARS-CoV and BtCoV-RaTG13. The high homology indicate that antibodies elicited by S1-76/77/78 may at least be effective for both SARS-Cov-2 and SARS-CoV.

These results strongly suggest that RBD is rich of conformational epitopes while poor of linear epitopes. The underlying mechanism is worth for further investigation. Taken together, our results suggest that S1-76/77/78 could serve as a promising candidate for the development of broadly neutralization antibodies or vaccine.

### CTD is rich of linear epitopes

The first “hot” region of linear epitopes is CTD. The whole domain is densely covered by linear epitopes (**Fig. 3A**). According to the selection criteria, 6 highly immunogenic epitopes, *i.e*., S1-93, S1-97, S1-105/106, S1-111 and S1-113 were identified. These peptides are about evenly distributed cross CTD. We then asked whether these 6 highly immunogenic peptides/ epitopes were also revealed by other studies. It showed that S1-93 was identified by ReScan (*23*)(**Table S3**) and COVIDep (*21*), S1-97 by ReScan, and S1-111 by COVIDep.

**Figure 3.**
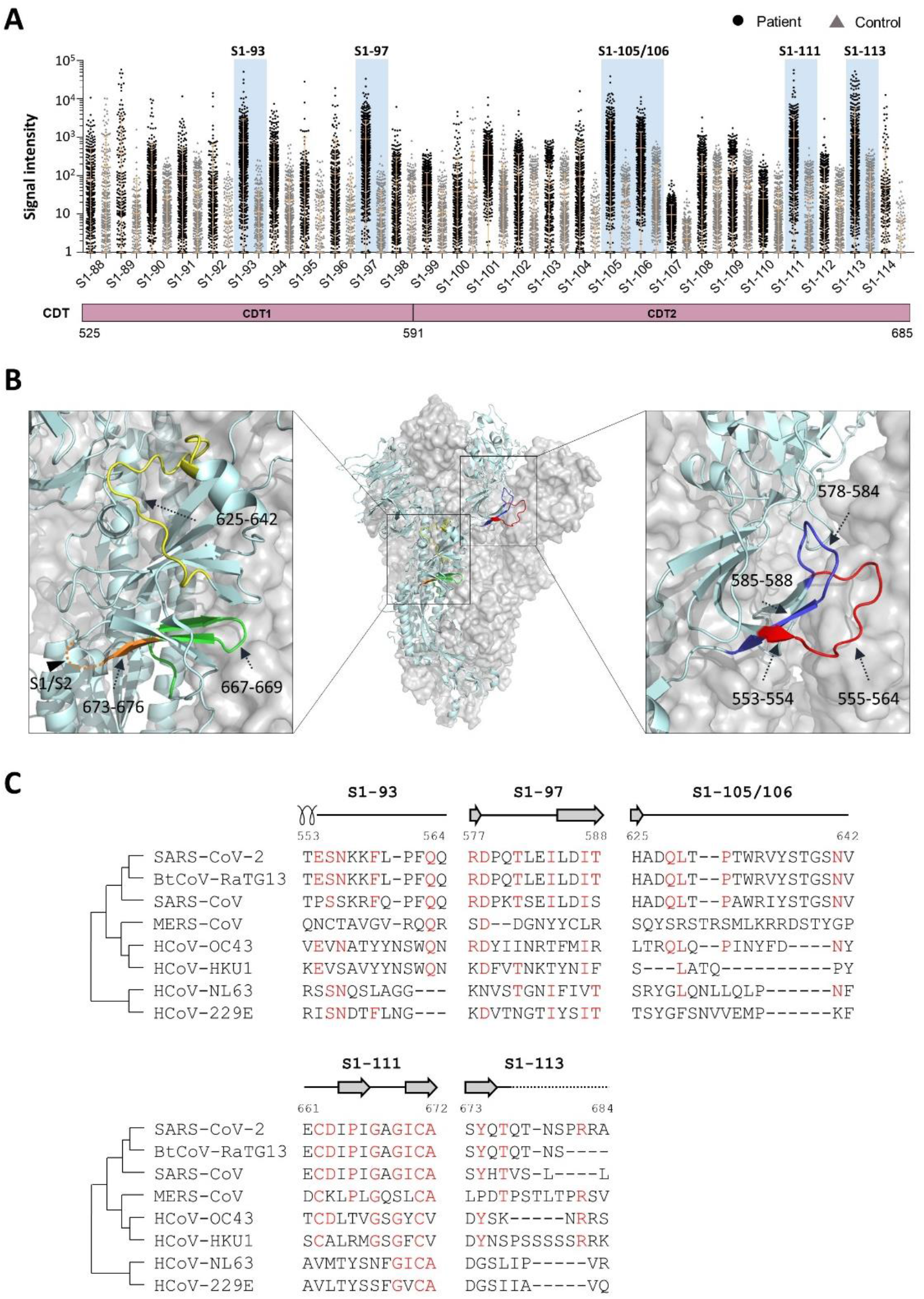
CTD is rich of significant linear epitopes. **A**. CTD region of the linear epitope landscape; **B**. The locations of the significant peptides locate on CTD (PDB ID: 6×6P). Specifically, S1-93, aa553-564, red; S1-97, aa577-588, blue; S1-105/106, aa625-642, yellow; S1-111, aa661-672, green; S1-113, S1-113, 673-684, orange; **C**. The homology analysis of the significant peptides among the 7 known human coronaviruses and the Bat coronavirus BtCoV-RaTG13. The amino acids with consistencies >=50% among the 8 coronaviruses are marked red, the loop, α-helix and β-strand region are shown as line, coil and arrow above sequences, respectively. Unobserved structure is shown as dotted line.

S1-93 and S1-97 locate at CTD1, aa555-564 of S1-93 and aa578-584 of S1-97 present as loop region and are on the surface of trimeric Spike protein. S1-105, S1-106, S1-111, S1-113 locate at CTD2, S1-105/106 are almost loop and present on the surface, S1-111 is β-strand and loop but buried inside, only aa667-669 on loop region could be accessed. S1-113 is nearly to S1/S2 cleavage site, although aa677-684 is invisible in Spike protein structure, these residues could be exposed on surface and induce antibody response to prevent S1/S2 hydrolysis (**Fig. 3B**). S1-113 is also on the out surface while S1-111 is on the inner surface, the underlying mechanism why S1-111 triggers strong IgG reaction in many patients worth further study.

Homology analysis was performed for the 6 highly immunogenic peptides (**Fig. 3C**). Except S1-113, high homologies were observed for all the 5 peptides among SARS-Cov-2, SARS-CoV and BtCoV-RaTG13. The high homology indicate that antibodies elicited by S1-93, S1-97, S1-105 and S1-111 may be effective for both SARS-Cov-2 and SARS-CoV. And antibody targeting S1-113 may specific for SARS-CoV-2.

D614G mutant is the current dominant strain in Europe (*36*), which has about 9 times higher infection efficiency in cell assay than that of the wild type strain (*37*). D614 is within S1-102, a peptide of moderate immunogenicity, and close to the highly immunogenic peptide S1-105. Block the D614 region may cause functionally significant effect.

### The second epitope hot region spans aa770-829, covers the S2’ cleavage site and FP

The second region with highly enriched linear epitopes spans aa770-829 (**Fig. 4A**). According to the selection criteria, 6 highly immunogenic peptides were obtained, *i.e*., S2-15, S2-16, S2-18, S2-19, S2-22 and S2-23.

**Figure 4.**
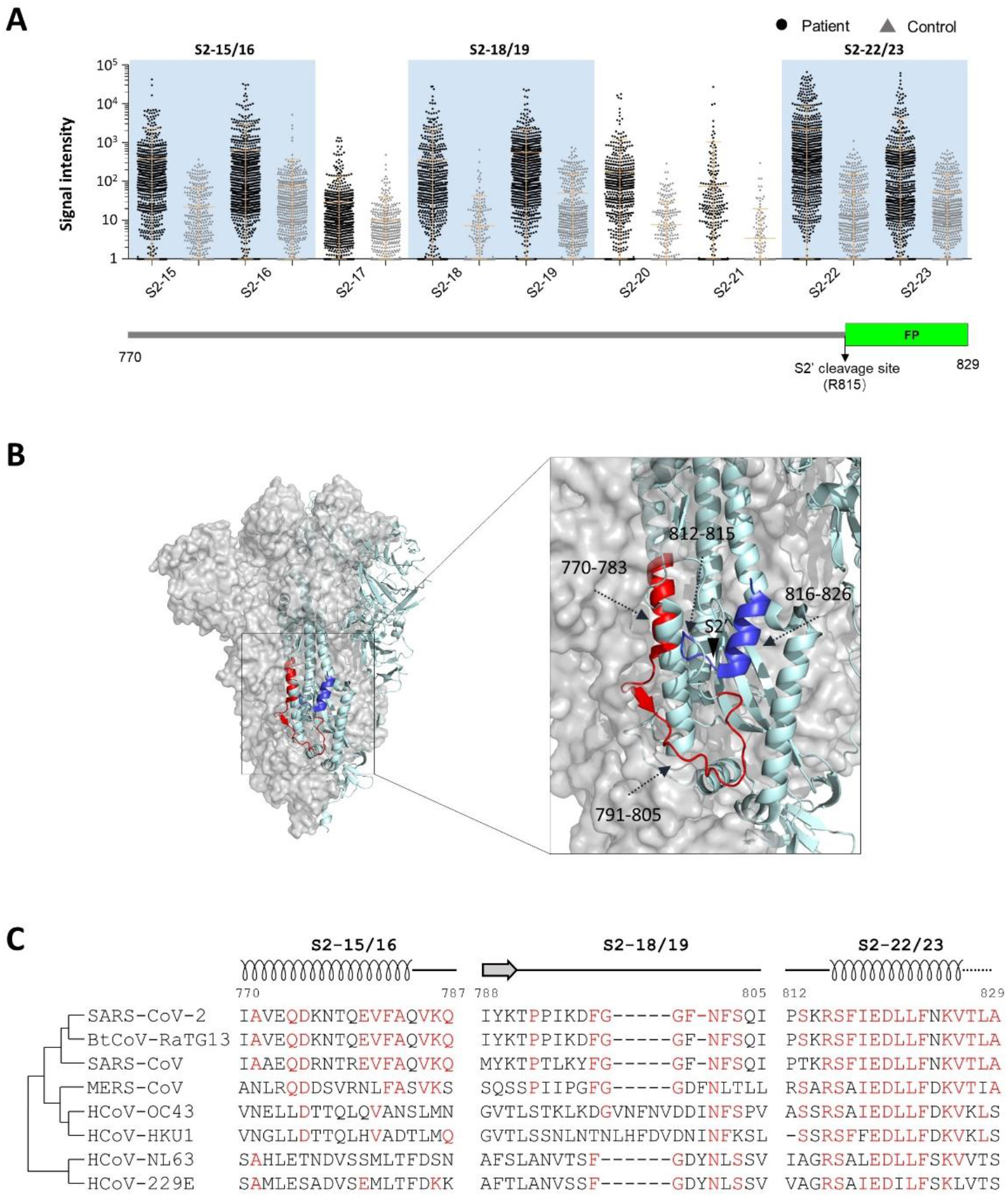
The 2^nd^ hot area of highly immunogenic linear epitopes: S2’cleavage site and FP. **A**. The S2’cleavage site and FP of the linear epitope landscape; **B**. The significant peptides locate on this region. S2-15/16, aa770-787, red, coil; S2-18/19, aa788-805, red, loop; S2-22/23, aa812-829, blue, coil; **C**. The homology analysis of the significant peptides among the 7 known human coronaviruses and the Bat coronavirus BtCoV-RaTG13. The amino acids with consistencies >=50% among the 8 coronaviruses are marked red. The loop, α-helix and β-strand region are shown as line, coil and arrow above sequences, respectively. Unobserved structure is shown as dotted line.

It is interesting to see whether these 6 highly immunogenic peptides/ epitopes were also identified in related studies. We revealed that S2-22 was identified by a peptide microarray study (*22*). Four peptides (S2-18, S2-19, S2-22 and S2-23) were identified by ReScan (*23*), and 2 peptides (S2-22 and S2-23) were predicted by COVIDep (*21*). Of these peptides, S2-22 is the only one that was identified or predicted in all these studies. Since S2-22/23 covers the S2’ cleavage site and FP, we speculate that antibody targeting S2-22/23 may block the cleavage and disturb the function of FP, thus has potent neutralization activity. Interestingly, strong S2-22 specific IgG reaction was also elicited by a mRNA vaccine study (*38*), which further demonstrated the high immunogenicity and high potential of neutralization activity of S2-22.

To further illustrate the locations of these peptides, we mapped them to the 3D structure of Spike protein. S2-15, S2-16, S2-18, S2-19, S2-22 and S2-23 all locate near S2’ site and the FP region. aa770-783 of S2-15/16 form as α-helix and are buried at the trimer interface, but accessible on monomer. aa791-805 of S2-18/19 form as loop and aa816-826 form as α-helix and locate on surface, the S2’ cleavage site is on S2-22 (**Fig. 4B**).

To check the similarity of the peptides among human coronaviruses, we performed the homology analysis for S2-22/23, S2-15/16 and S2-18/19. High homologies were observed for all these peptides among SARS-Cov-2, SARS-CoV and BtCoV-RaTG13 (**Fig. 4C**). Interestingly, S2-22/23 is highly homologous among all the coronaviruses, and almost identical among SARS-Cov-2, SARS-CoV and BtCoV-RaTG13. If this peptide can elicit strong neutralization activity, it may could serve as a promising candidate for making broad neutralization antibody and vaccine.

### Other highly immunogenic linear epitopes

Except the immunogenic peptides that belong to RBD and the two “hot” regions, there are also other highly immunogenic peptides dispersed across Spike protein (**Fig. 5A**). S1-5 locates at NTD, part of the residues, *i. e*., aa28-31 form β-strand and are on the surface of trimeric Spike protein, aa32-36 form as loop and are partially covered by other region. S2-78, S2-96 and S2-97 locate at unobserved regions in the C-terminal of Spike protein. We applied a modeling structure to present these unobserved regions (**Fig. 5B**). S2-78 is predicted as α-helix and S2-96/97 is predicted as loop. S2-96/97 are at the very C-terminal end of Spike protein (**Fig. 5B**), which locates in the cytoplasm of the host cell.

**Figure 5.**
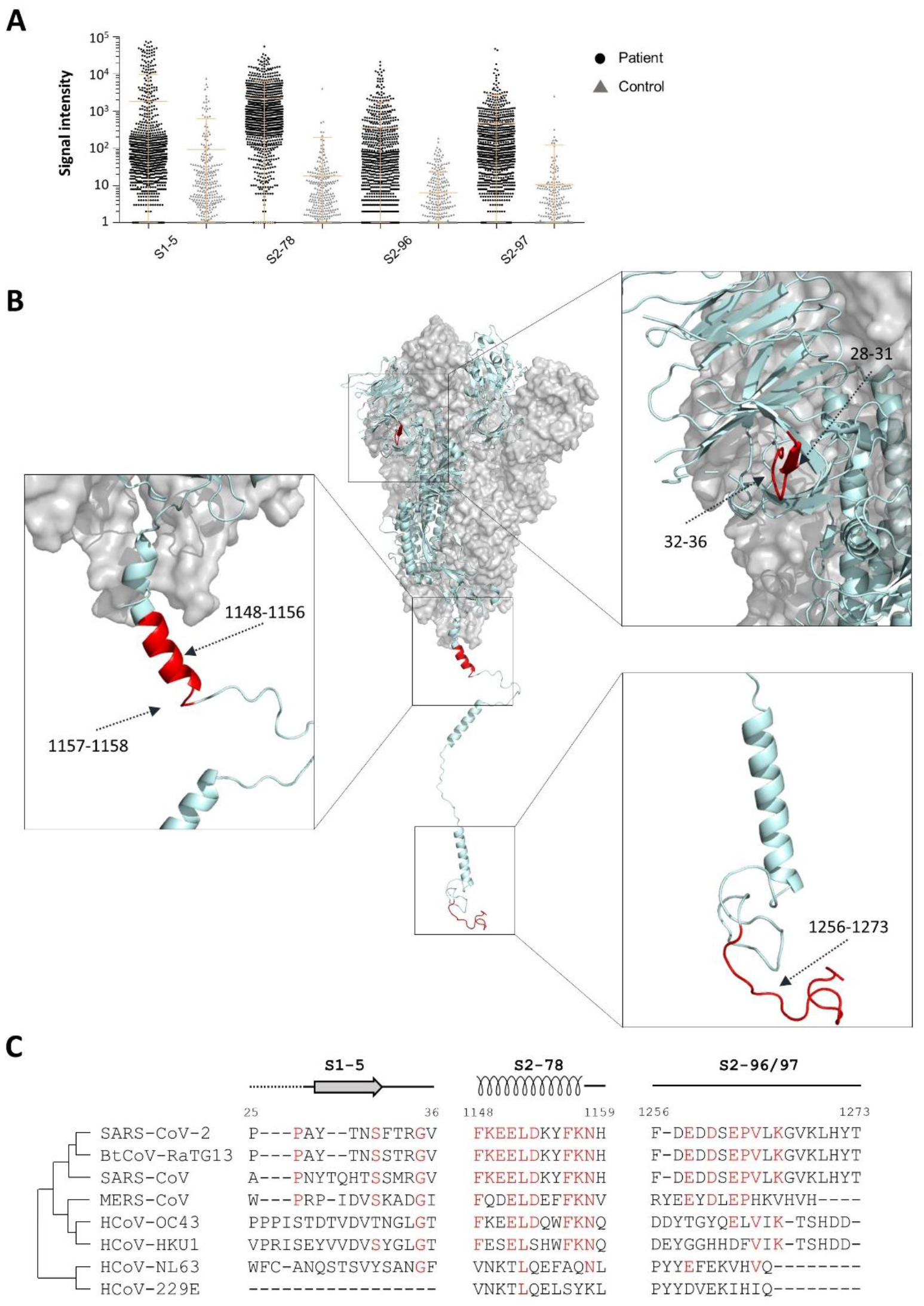
Other highly immunogenic linear epitopes. **A**. Other 5 significant peptides which are not belong the two “hot regions” (see **Figure 3** and **Figure 4**); **B**. The significant peptides locate on Spike protein. S1-5, aa25-36, red; S2-78, aa1148-1159, red; S2-96/97, aa1256-1273, red; **C**. The homology analysis of the significant peptides among the 7 known human coronaviruses and the Bat coronavirus BtCoV-RaTG13. The amino acids with consistencies >=50% among the 8 coronaviruses are marked red. The loop, α-helix and β-strand region are shown as line, coil and arrow above sequences, respectively. Unobserved structure is shown as dotted line.

We checked whether these 4 highly immunogenic peptides/ epitopes were also revealed in other studies. It showed that S1-5 was identified by a peptide microarray study (*22*). S2-78 was identified by ReScan (*23*), and 3 peptides (S2-78, S2-96, S2-97) were predicted by COVIDep (*21*). The functional role of S1-5 specific antibodies may worth further investigation. S2-78 is adjacent to HR2, the antibody targeting this site may block the conformational change that is essential for effective virus-cell fusion (*39*). It is surprising to see the high immunogenicity of S2-96 and S2-97, since they are at the very C-terminal end of Spike protein and theoretically localize in the cytoplasm. Further study is necessary to explore the underlying mechanism and functional roles of these peptides/ epitopes.

High homologies were observed for all these peptides among SARS-Cov-2, SARS-CoV and BtCoV-RaTG13 (**Fig. 5C**). Interestingly, S2-78 and S2-96/97 are highly homologous among all the coronaviruses, and almost identical among SARS-Cov-2, SARS-CoV and BtCoV-RaTG13. For S2-78, high homology was also observed for MERS-CoV. Same as S2-22/23, S2-78 has the potential to serve as a promising candidate for developing broad neutralization antibody and vaccine.

## Discussion

Herein, we aim to reveal IgG responses triggered by SARS-CoV-2 spike protein on a systematic level. We adopted a newly developed peptide microarray with full coverage of Spike protein (*14*), analyzed 1,051 COVID-19 sera and 528 control sera. A set of highly immunogenic peptides/epitopes were revealed, and a comprehensive IgG linear epitope landscape was constructed.

One limitation of this study is that only short peptides were involved. Though linear epitopes are nicely represented, conformational epitopes may not, for example, for RBD region, which is highly immunogenic, only 3 linear epitopes of moderate immunogenicity were identified. To overcome this limitation, one way is to synthesize longer peptides which may retain some conformational information (*31*). It is necessary to point out that for the linear epitopes that we identified, they are highly physiologically relevant, since all of them were revealed through the analysis of sera from COVID-19 patient.

Our study presents the first IgG linear epitope landscape of Spike protein, which could only be enabled by analyzing a large cohort of samples using a systematic approach, such as the peptide microarray of full coverage of Spike protein. According to the landscape, it is obvious that Spike protein is highly immunogenic, there are many epitopes on the protein, and these epitopes are not evenly distributed across Spike protein. Among the 19 immunogenic peptides (**Table S3**), some may be good candidates for developing neutralization antibodies or vaccines, some may cause ADE, if any. The rest of the peptides may have no direct biological function, but serve only as neutral immunodominant epitopes that could not trigger neutralization activity and ADE. We believe that most of the 19 immunogenic peptides (**Table S3**) are worth further testing in animal model to identify which peptides can specifically trigger neutralization activities. When use the intact Spike protein for vaccine development, we can take a more precise way by deleting or mutating the peptides that correspond to ADE and neutral immunodominant epitopes, thus assure the efficacy while minimize the side effect. Alternatively, the peptides which elicit neutralization activity have the potential to be applied directly for neutralization antibody generation, or applied as peptide vaccine. By this way, we can avoid many challenges when working with intact Spike protein or RBD.

The linear epitope landscape and the highly immunogenic peptides identified in this study could serve as a solid base to guide the precise development of prophylactic/ therapeutic antibodies and vaccines for combating COVID-19. Because of the high homology, they may also effective for other human coronaviruses, including SARS-CoV and MERS-CoV. Since most of the highly immunogenic peptides are outside of RBD, we believe that the antibodies and vaccines based on these peptides/epitopes could be used alone or at least as a complementary to the RBD centered antibodies and vaccines.

## Materials and Methods

### Peptide synthesis and conjugation with BSA

The N-terminal amidated peptides were synthesized by GL Biochem, Ltd. (Shanghai, China). Each peptide was individually conjugated with BSA using Sulfo-SMCC (Thermo Fisher Scientific, MA, USA) according to the manufacture’s instruction. Briefly, BSA was activated by Sulfo-SMCC in a molar ratio of 1: 30, followed by dialysis in PBS buffer. The peptide with cysteine was added in a w/w ratio of 1:1 and incubated for 2 h, followed by dialysis in PBS to remove free peptides. A few conjugates were randomly selected for examination by SDS-PAGE. For the conjugates of biotin-BSA-peptide, before conjugation, BSA was labelled with biotin by using NHS-LC-Biotin reagent (Thermo Fisher Scientific, MA, USA) with a molar ratio of 1: 5, and then activated by Sulfo-SMCC.

### Peptide microarray fabrication

The peptide-BSA conjugates as well as S1 protein, RBD protein and N protein of SARS-CoV-2, along with the negative (BSA) and positive controls (anti-Human IgG and IgM antibody), were printed in triplicate on PATH substrate slide (Grace Bio-Labs, Oregon, USA) to generate identical arrays in a 1 x 7 subarray format using Super Marathon printer (Arrayjet, UK). The microarrays were stored at −80°C until use.

### Patients and samples

The study was approved by the Ethical Committee of Tongji Hospital, Tongji Medical College, Huazhong University of Science and Technology, Wuhan, China (IRB ID:TJ-C20200128) Written informed consent was obtained from all participants enrolled in this study. COVID-19 patients were hospitalized and received treatment in Tongji Hospital during the period from 17 February 2020 and 28 April 2020. Serum from each patient was collected when hospitalization at viable time points (**Table S1**). Sera of the control group from healthy donors, lung cancer patients, patients with autoimmune diseases were collected from Ruijin Hospital, Shanghai, China or Tongren Hospital, Shanghai, China. All the sera were stored at −80°C until use.

### Microarray-based serum analysis

A 14-chamber rubber gasket was mounted onto each slide to create individual chambers for the 14 identical subarrays. The microarray was used for serum profiling as described previously with minor modifications(*40*). Briefly, the arrays stored at −80°C were warmed to room temperature and then incubated in blocking buffer (3% BSA in 1×PBS buffer with 0.1% Tween 20) for 3 h. A total of 200 μL of diluted sera or antibodies was incubated with each subarray for 2 h. The sera were diluted at 1:200 for most samples and for competition experiment, free peptides were added at a concentration of 0.25 mg/mL. For the enriched antibodies, 0.1-0.5 μg antibodies were included in 200 μL incubation buffer. The arrays were washed with 1×PBST and bound antibodies were detected by incubating with Cy3 - conjugated goat anti-human IgG and Alexa Fluor 647-conjugated donkey anti-human IgM (Jackson ImmunoResearch, PA, USA), which were diluted for 1: 1,000 in 1×PBST. The incubation was carried out at room temperature for 1 h. The microarrays were then washed with 1×PBST and dried by centrifugation at room temperature and scanned by LuxScan 10K-A (CapitalBio Corporation, Beijing, China) with the parameters set as 95% laser power/ PMT 550 and 95% laser power/ PMT 480 for IgM and IgG, respectively. The fluorescent intensity was extracted by GenePix Pro 6.0 software (Molecular Devices, CA, USA).

### Data analysis of peptide microarray

For each spot, signal intensity was defined as the mean_foreground subtracted by the mean_background. The signal intensities of the triplicate spots for each peptide or protein were averaged. The overall_mean_background and the overall_standard deviation (SD)_background of all the arrays probed with COVID-19 sera were calculated. Cutoff1 was defined as (the overall_mean_background + 2*overall_SD_background). According to the array data, Cutoff1 was calculated as 380.7. For the control arrays, mean_forground and SD_foreground for each peptide and protein were calculated. Cutoff2 was set as (control_mean_singal intensity + 2*control_SD_ signal intentisy). For each peptide or protein, SARS-CoV-2 specific positive response was called when the average_signal intesnty is larger than both Cutoff1 and Cutoff2. Response frequency was then defined as the number of the peptides with positive response divided by the total number of the peptides on the microarray.

### Structure analysis

The spike protein structures (PDB ID: 6×6P and 6VYB), RBD-ACE2 structure (PDB ID: 6M0J) and antibodies-RBD complex structure (PDB ID: 7C01, 7BWJ, 7BYR, 6W41 and 6WPT) were used to analyze the structural details of the epitopes identified from the peptide microarray. The C terminal (1146-1273) structure of Spike protein was from a modeling structure, QHD43416.pdb, generated by the C-I-TASSER (https://zhanglab.ccmb.med.umich.edu/COVID-19/), and aligned to C terminal of Spike protein (PDB ID: 6×6P). Structural analysis was processed in Pymol. The alignment and homology analysis of 7 human coronaviruses and one bat coronavirus was generated by ClustalW algorithm from EMBL-EBI (https://www.ebi.ac.uk/Tools/msa/clustalo/).

## Data Availability

The peptide microarray data are deposited on Protein Microarray Database (http://www.proteinmicroarray.cn)

## Acknowledgments

We thank Dr. Daniel M. Czajkowsky for critical reading and editing.

## Author contributions

S-C. T. developed the conceptual ideas and designed the study. Z-Y. S., F.W., H-Y. H., Y-D. Z., X-S. L., Z-J. Y., H-M. S. and J-X. W. collected the sera samples. X-L F., Y. L., M-L. M., Z-W. X., B. Z., H. C., C-Z. Y., J-B. X., X-N. W., Y-X. Z., D-Y. L., H-N. Z., H-W. J., H. Q., and S-J. G. performed the experiments. S-C.T., Y. L. and M-L. M. wrote the manuscript with suggestions from other authors.

## Data and materials availability

Additional data related to this paper may be requested from the authors.

---

**Figure S1.**
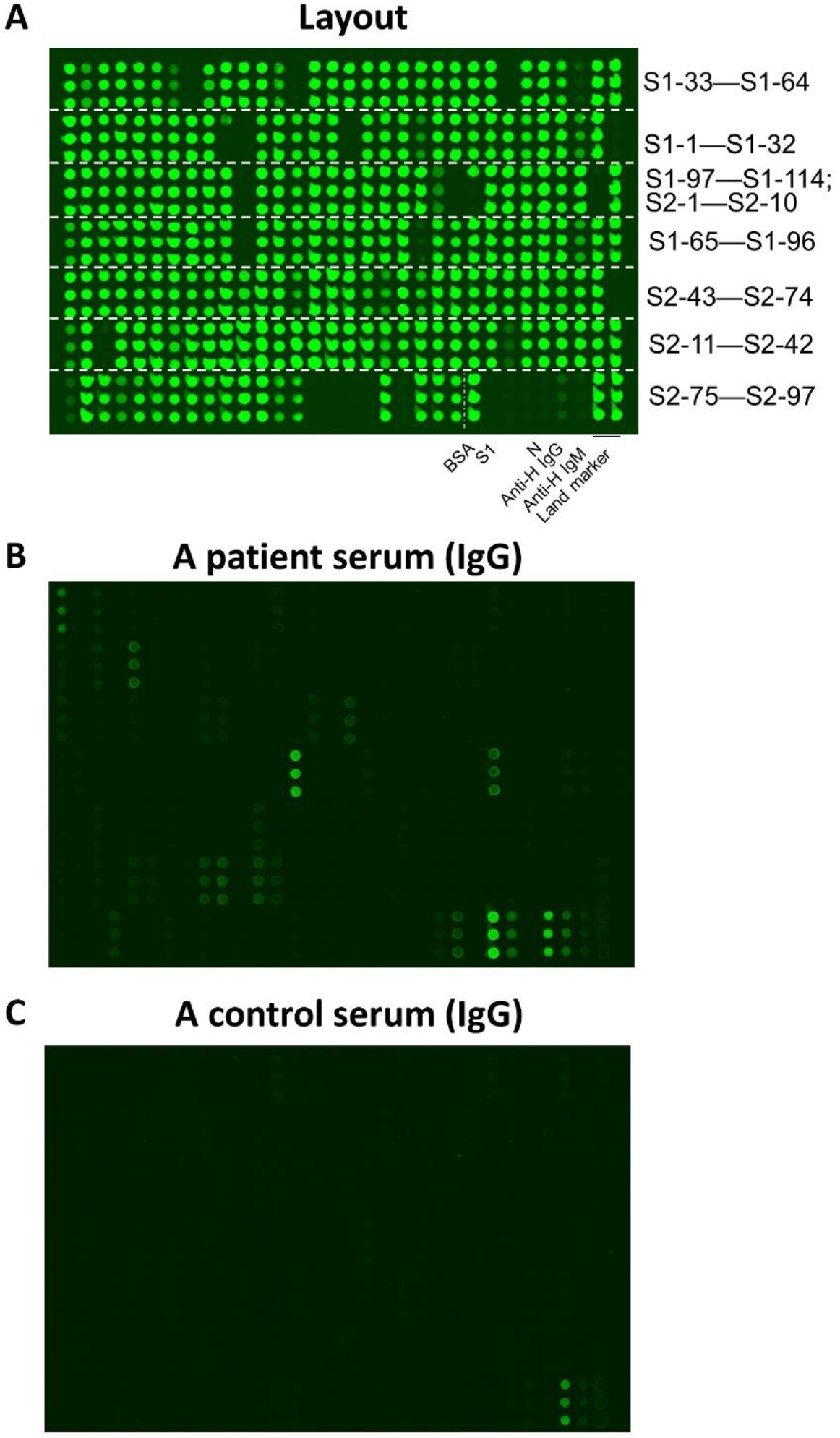
The peptide microarray. **A**. The layout of the peptide microarray that was used in this study (Please see **Table S2** for the details of the peptides on the array); **B**. An example array probed with COVID-19 patient serum; **C**. An example array probed with a control serum.

**Figure S2.**
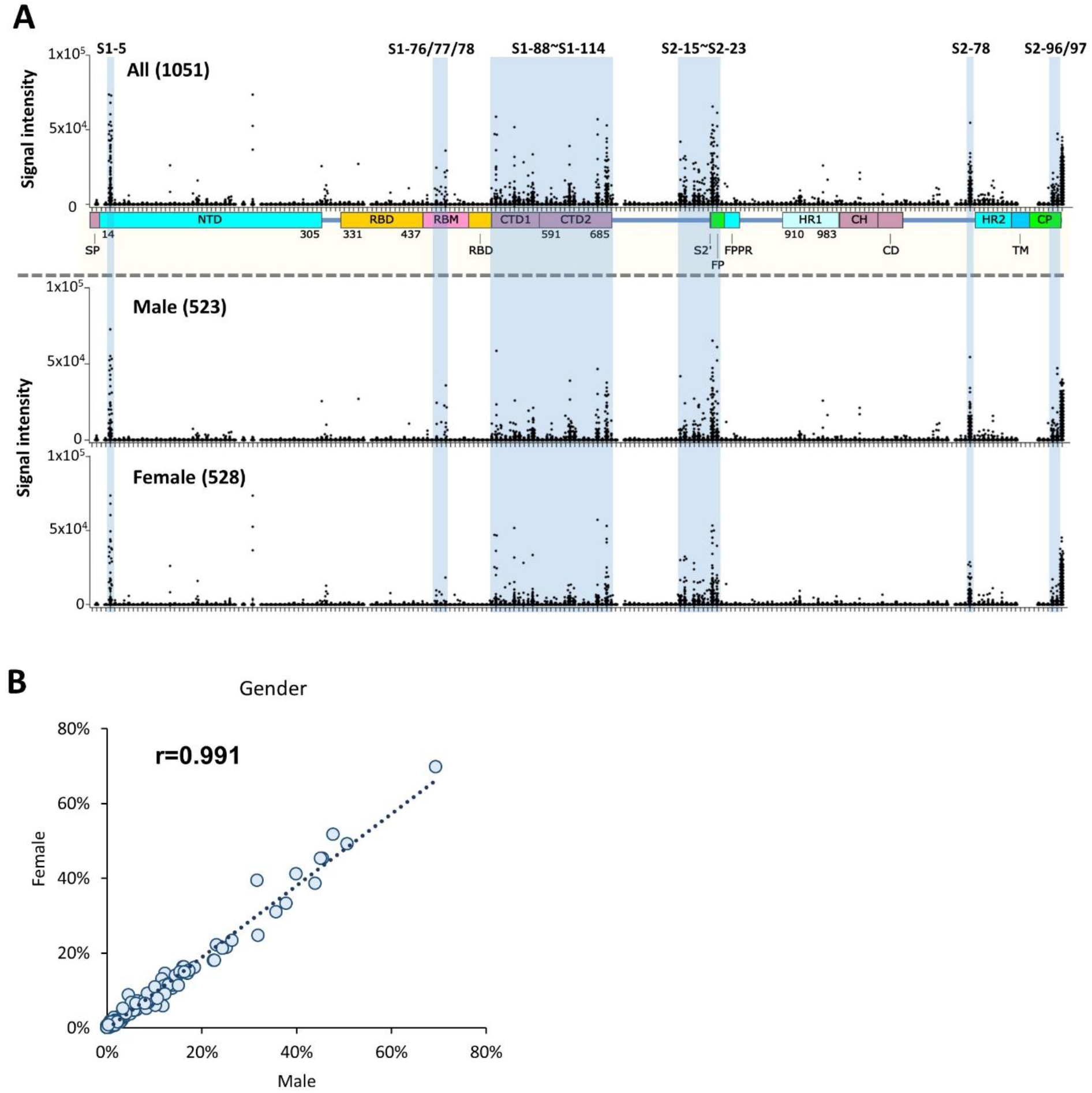
The IgG linear epitope landscape of SARS-CoV-2 Spike protein: male vs. female. **A**. The linear epitope landscape was divided into two sub-landscapes according to gender, *i.e*., 523 male vs. 528 female; **B**. The correlation between male and female.

**Figure S3.**
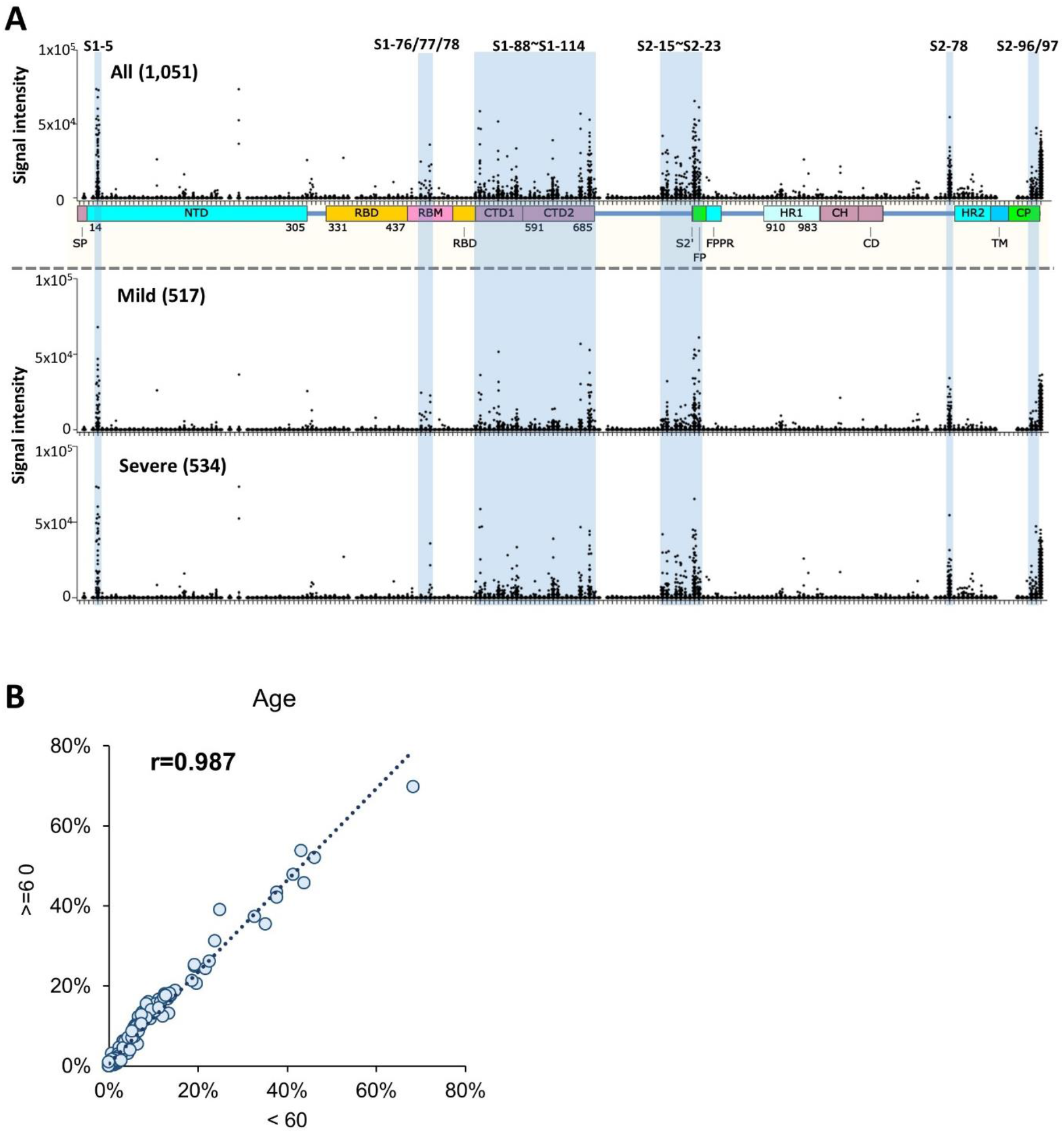
The IgG linear epitope landscape of SARS-CoV-2 Spike protein: mild vs. severe. **A**. The linear epitope landscape was divided into two sub-landscapes according to severity, *i.e*., 513 mild vs 534 severe (including severe and critical cases); **B**. The correlation between mild and severe.

**Figure S4.**
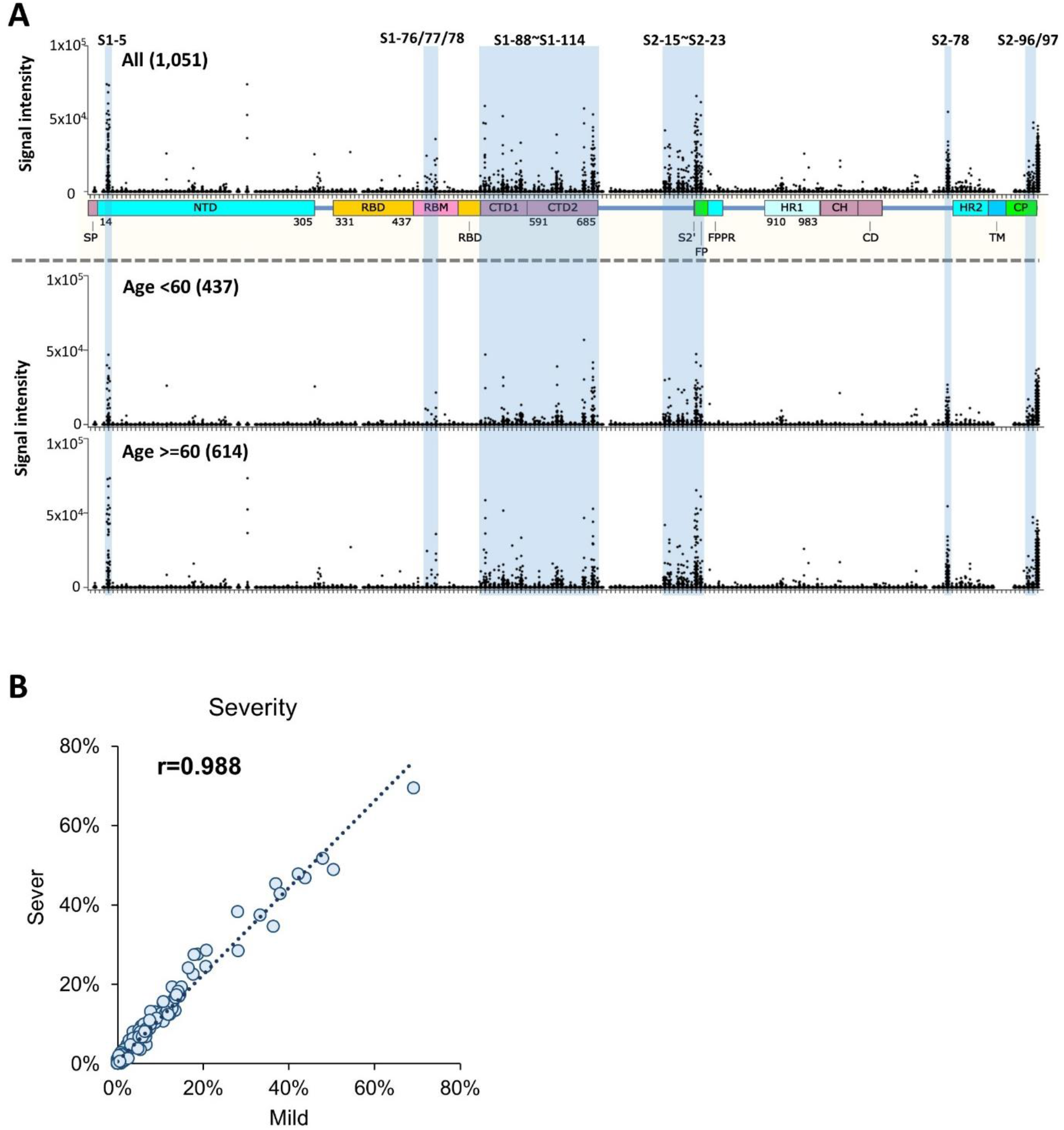
The IgG linear epitope landscape of SARS-CoV-2 Spike protein: age < 60 vs. >= 60. **A**. The linear epitope landscape was separated to two sub-landscapes according to age, *i.e*., 437 age < 60 vs. 614 age >= 60; **B**. The correlation between age >=60 and age < 60.

**Figure S5.**
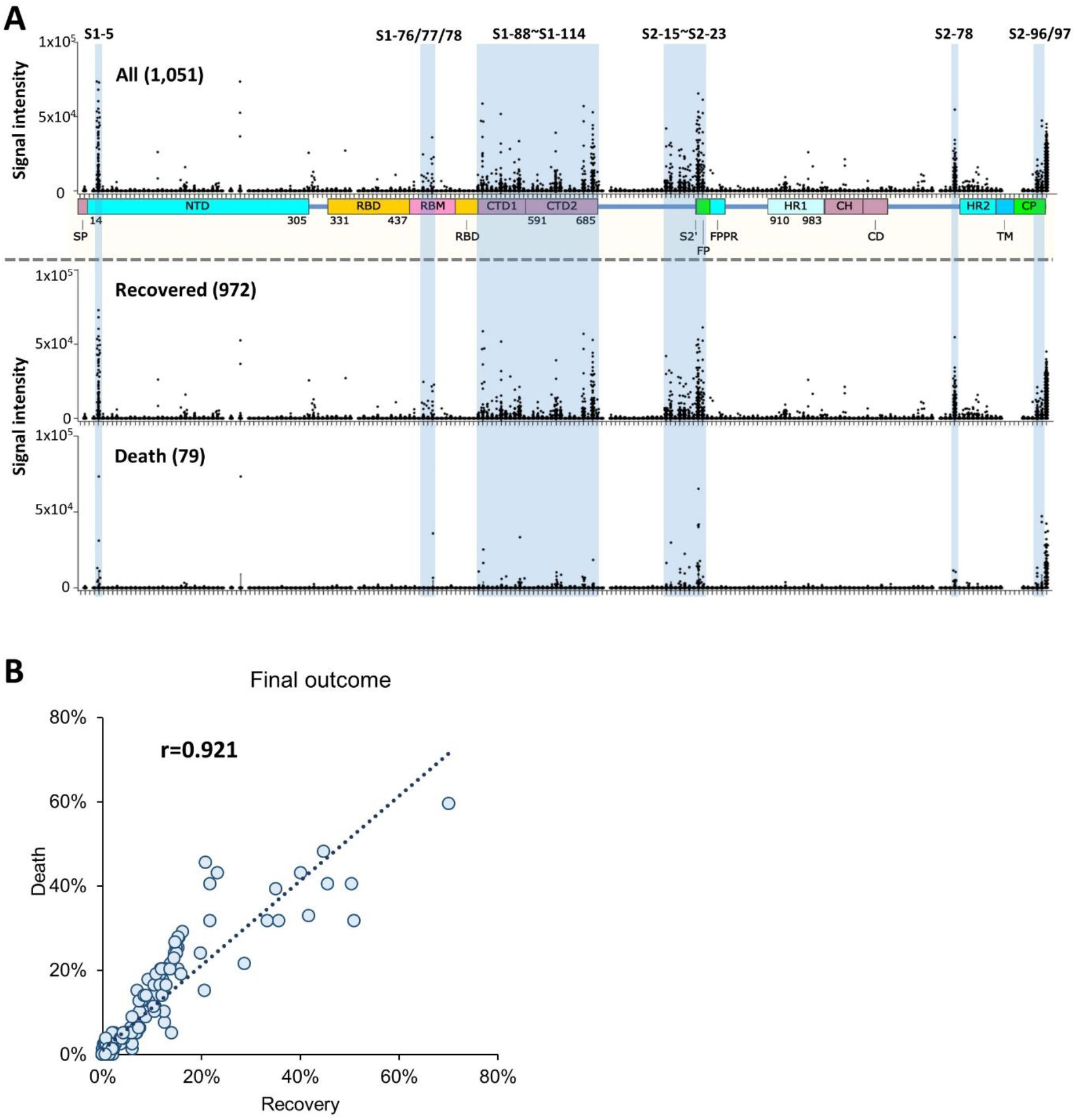
The IgG linear epitope landscape of SARS-CoV-2 Spike protein: recovered vs. death. **A**. The linear epitope landscape was divided into two sub-landscapes according to the final outcome, *i.e*., recovered (972) vs. death (79); **B**. The correlation between recovered and death.

**Figure S6.**
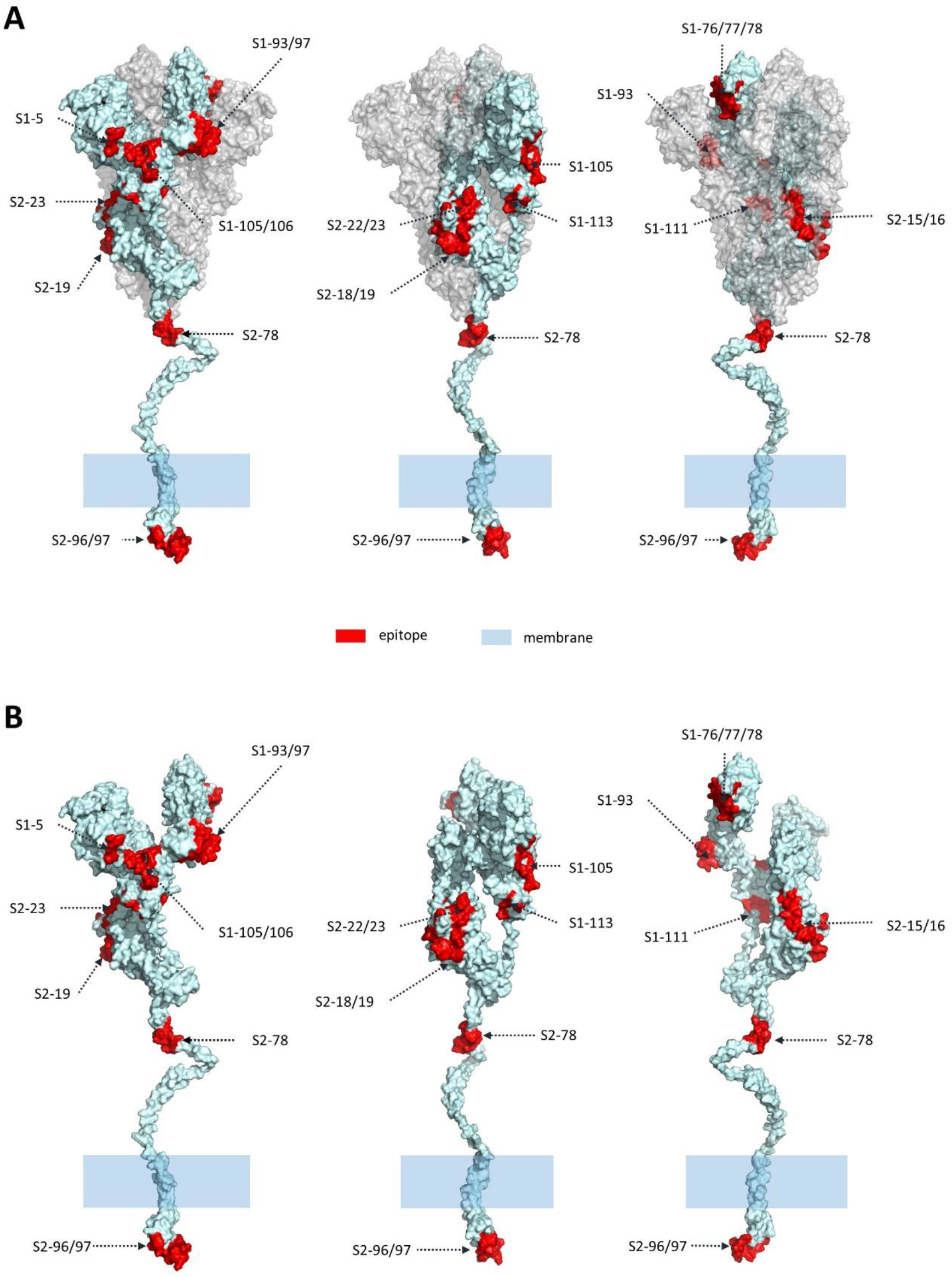
The distribution of the highly immunogenic peptides on Spike protein. A 3D structure of spike protein is adopted (PDB ID: 6×6P). The part from S2-78 to the end of C-terminal was modeled by C-I-TASSER. The 19 significant peptides are red marked on the 3D structure of Spike protein for both trimer (**A**) and monomer (**B**).

**Figure S7.**
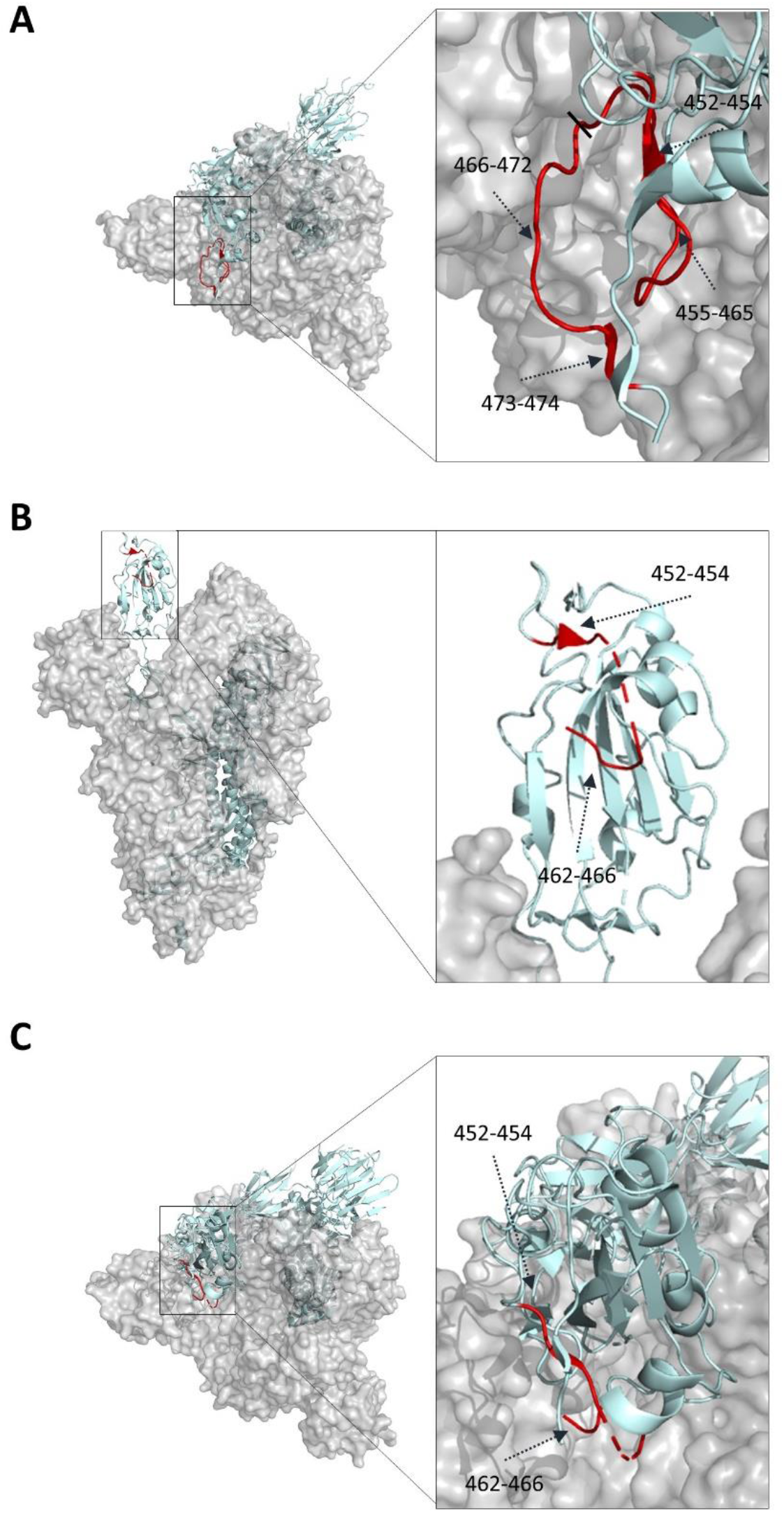
The location of S1-76/77/78 on RBD. **A**. The top view of closed state Spike protein trimer (PDB: 6×6P); **B**. The side view of open state Spike protein trimer (PDB ID: 6VYB); **C**. The top view of open state Spike protein trimer (PDB ID: 6VYB). The significant peptides (S1-76/77/78, aa451-474) were marked as red.

**Figure S8.**
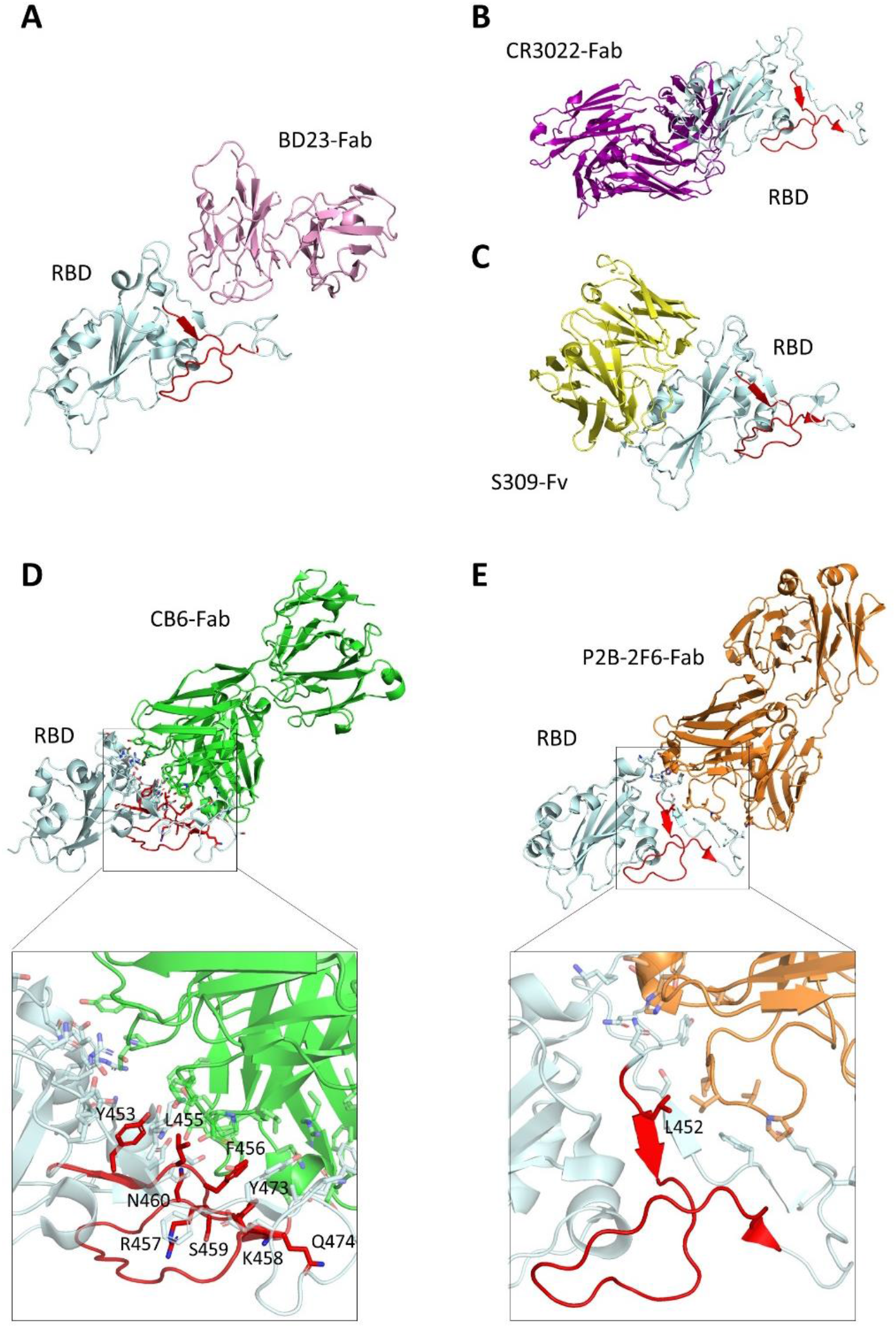
The overlap of S1-76/77/78 with neutralization antibodies. **A**. RBD-BD23-Fab complex (PDB: 7BYR); **B**. RBD-CR3022-Fab complex (PDB ID: 6W41); **C**. RBD-S309-Fv complex (PDB ID: 6WPT); **D**. RBD-CB6-Fab complex (PDB ID: 7C01), a close-up of the interface between RBD and CB6-Fab is depicted below, the side chain of contacting residues is shown as stick; **E**. RBD-P2B-2F6-Fab complex (PDB ID: 7BYR), a close-up of the interface between RBD and P2B-2F6-Fab is depicted below, the side chain of contacting residues are shown as stick. The significant peptides (S1-76/77/78, aa451-474) were marked as red (**A-E**).

**Table. S1.**
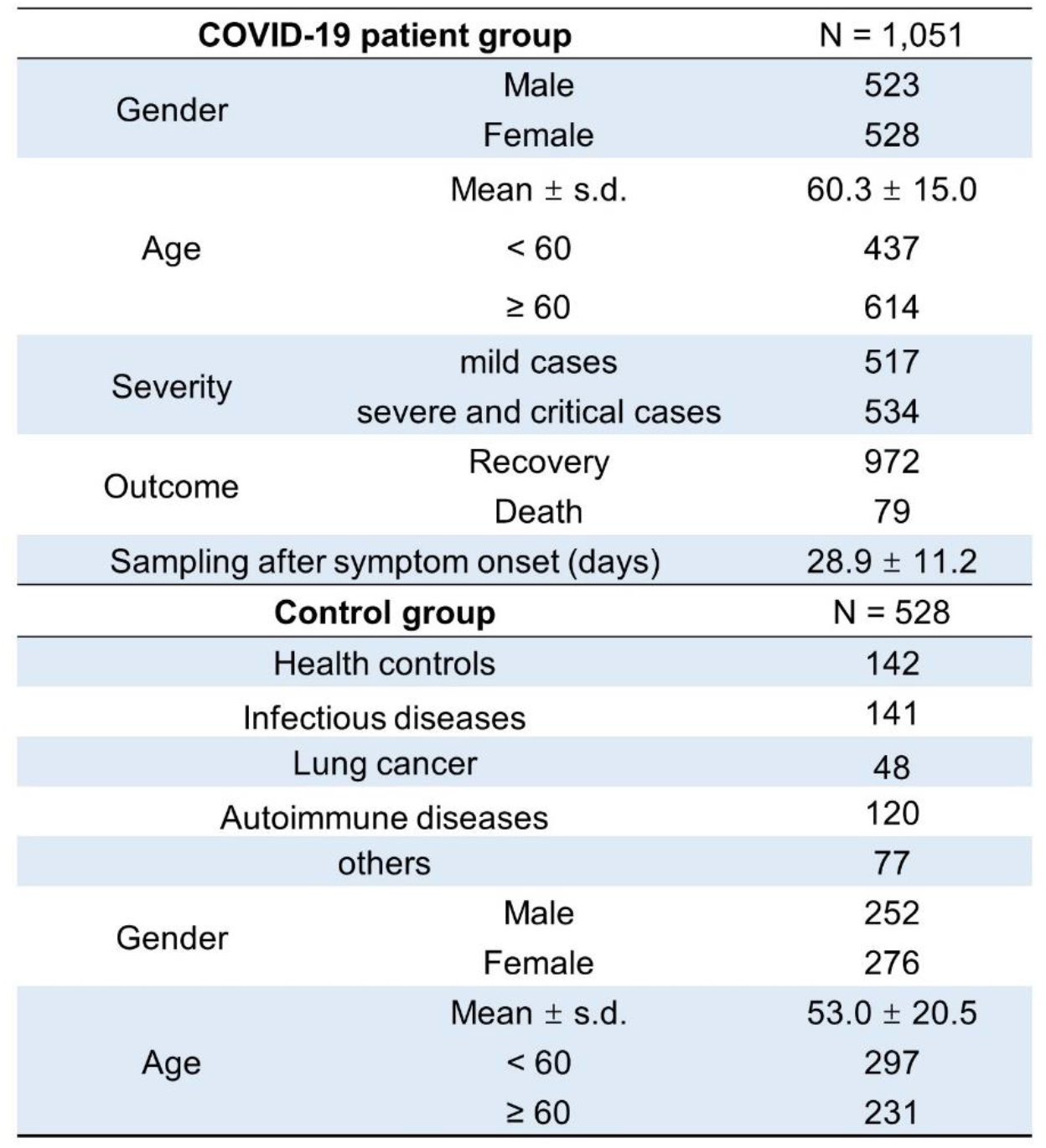
Serum samples tested in this study.

**Table S2.**
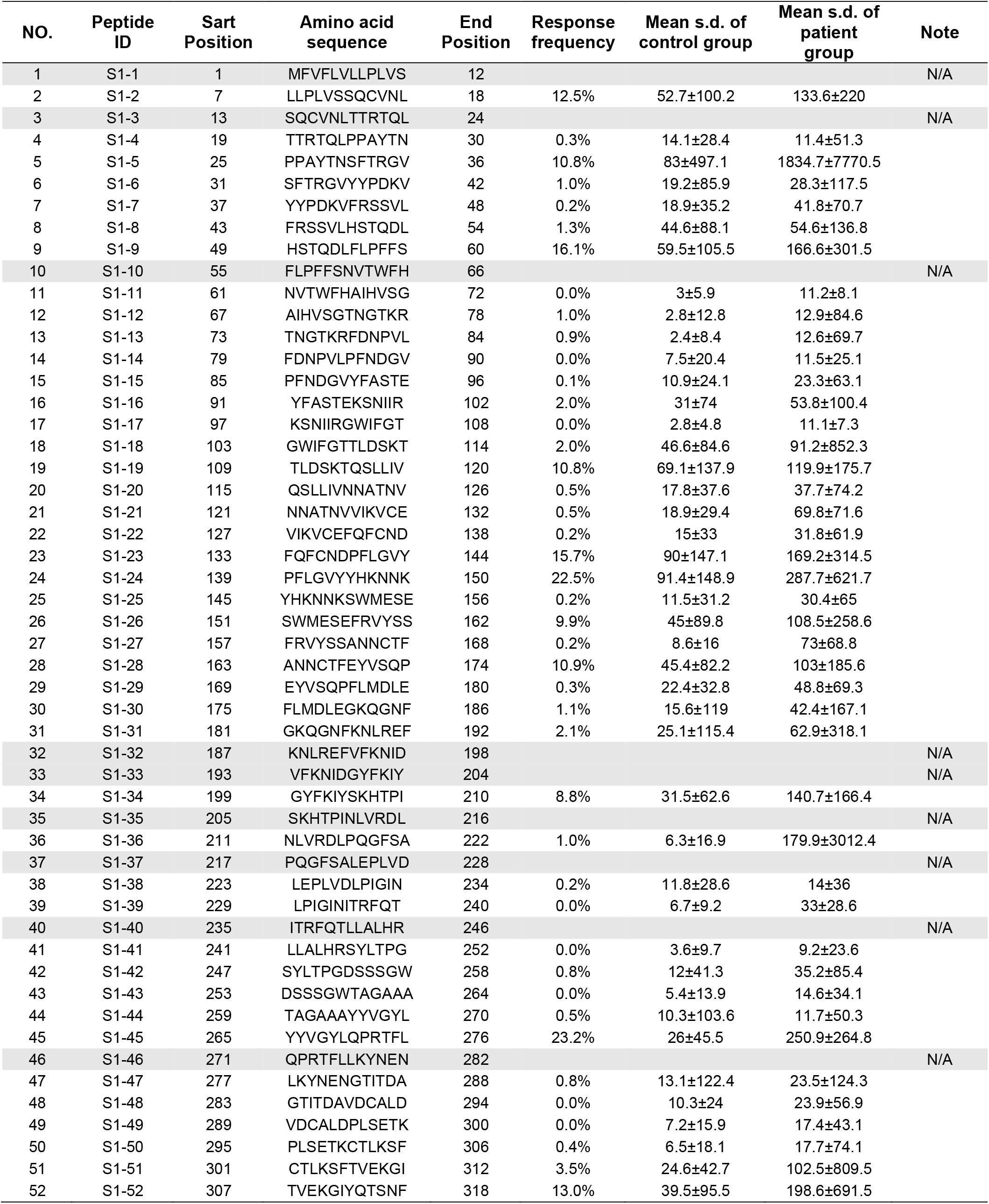
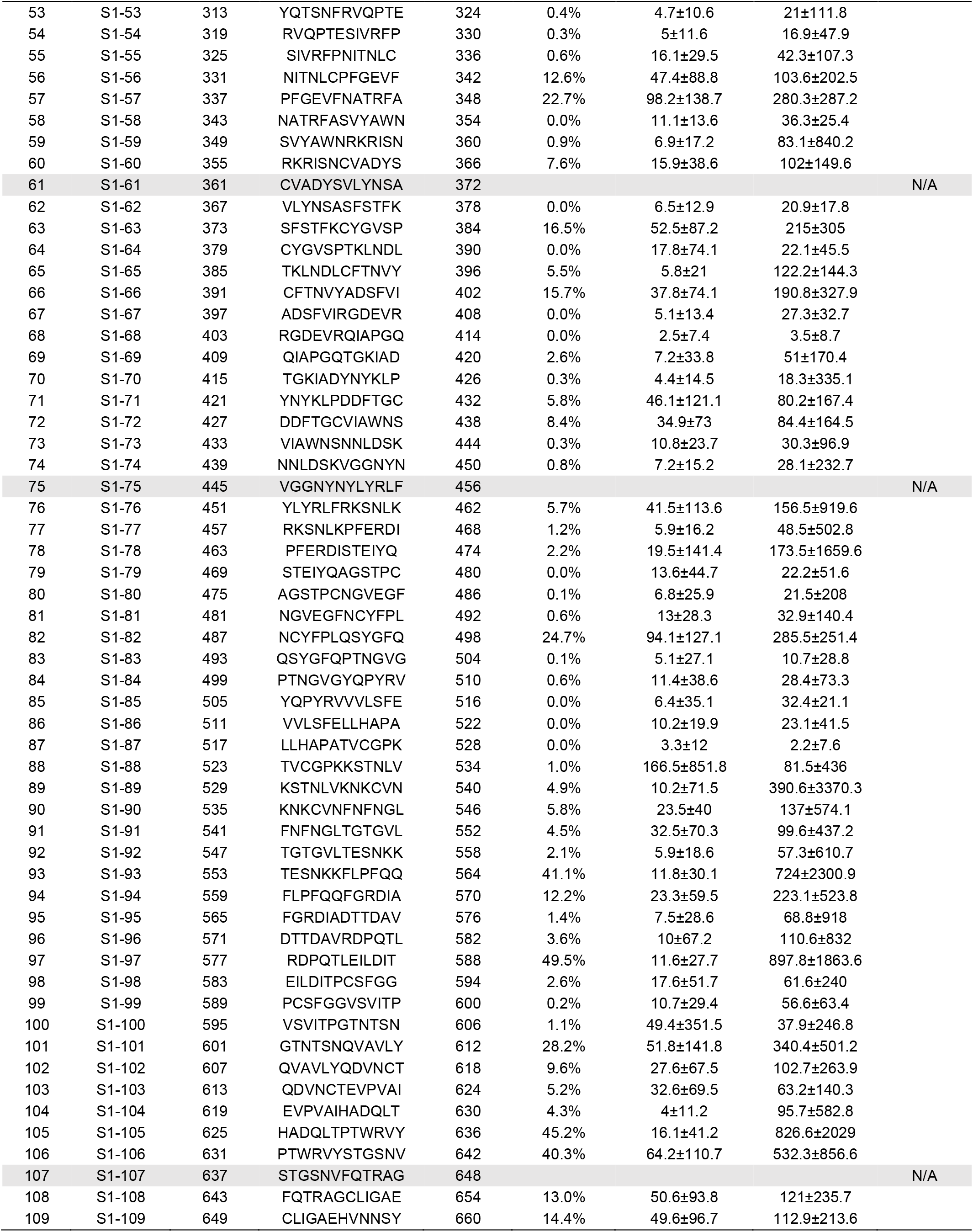
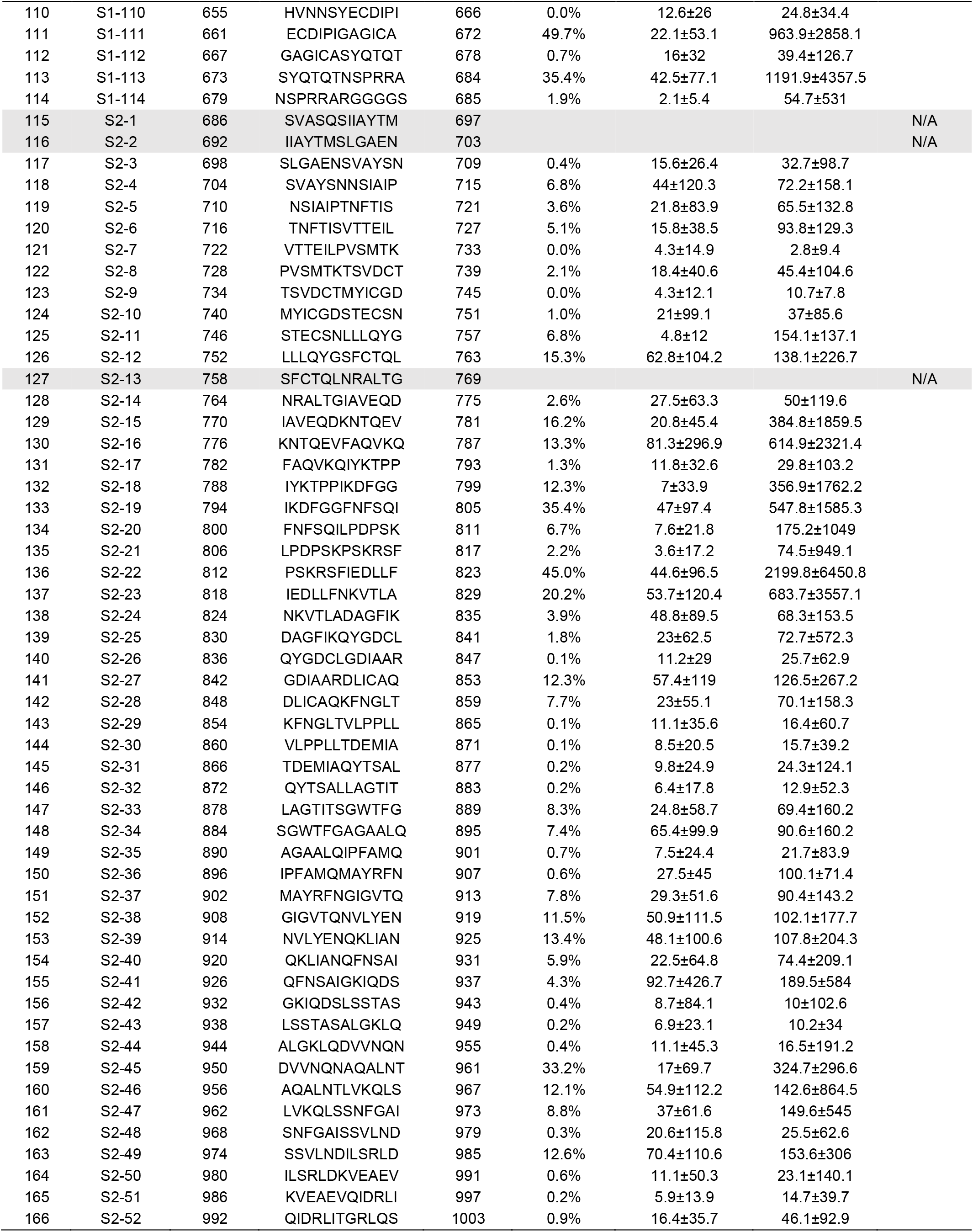
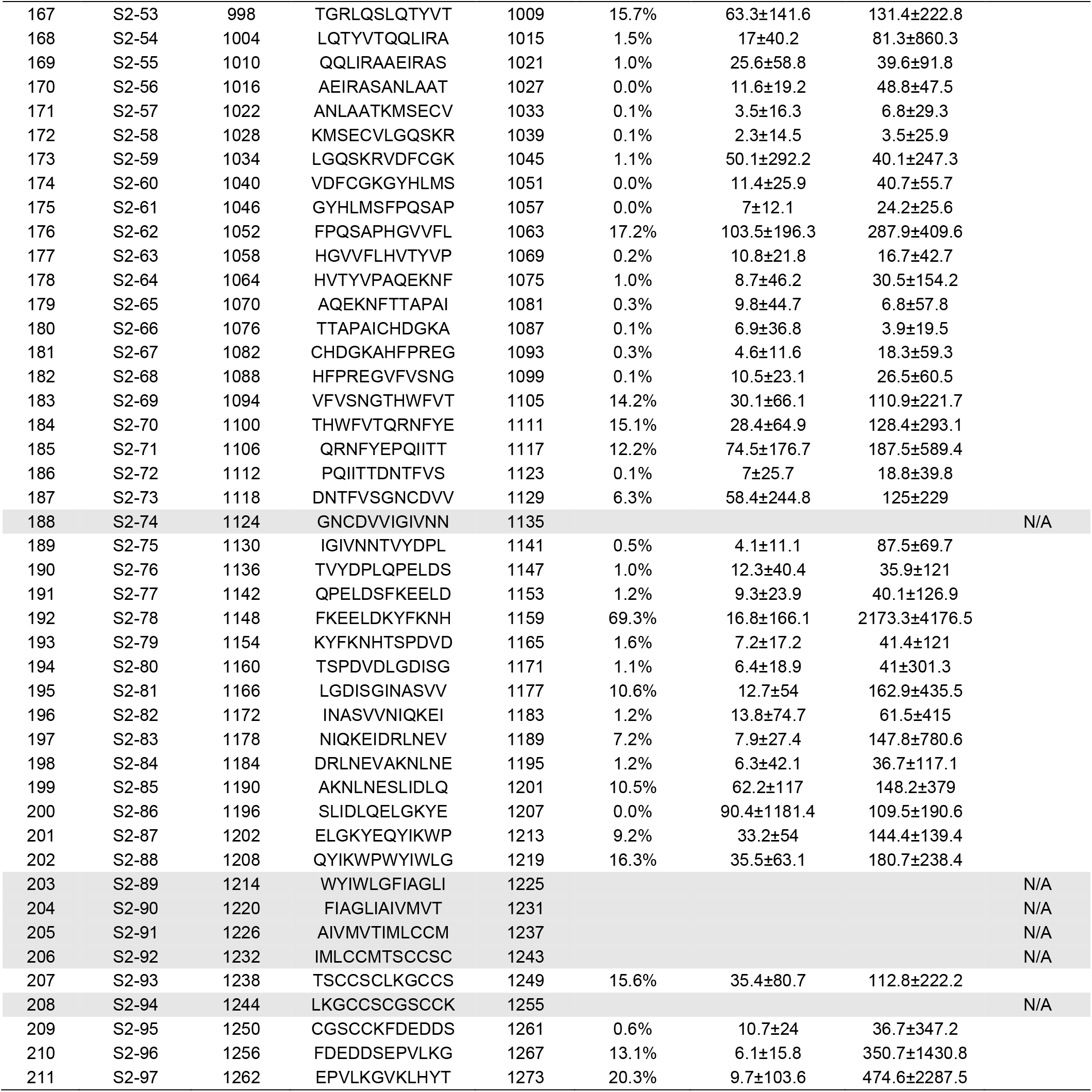
Highly immunogenic peptides.

**Table S3.**
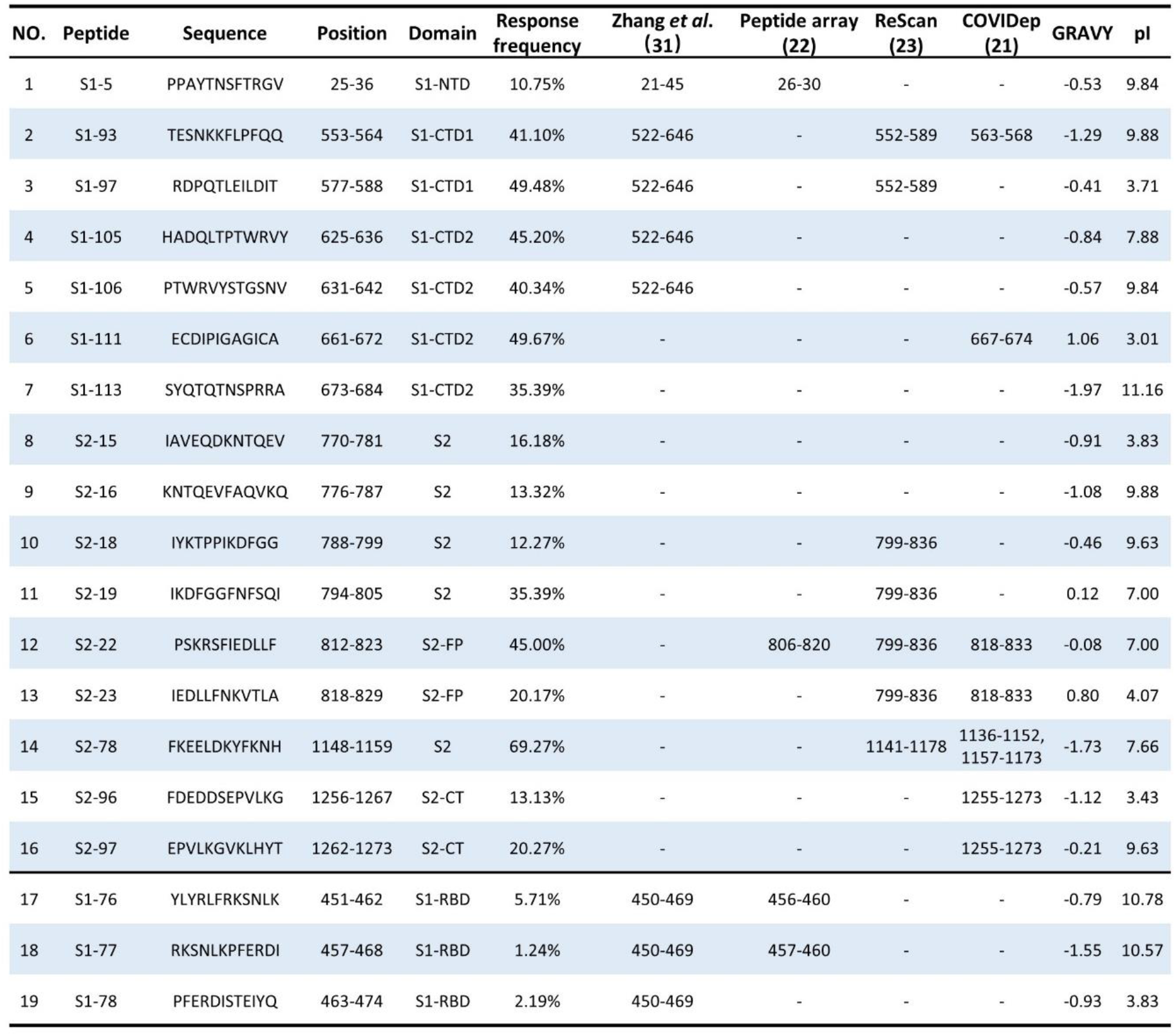
Highly immunogenic peptides.

